# Monocyte CD169 expression as a biomarker in the early diagnosis of COVID-19

**DOI:** 10.1101/2020.06.28.20141556

**Authors:** Anne-Sophie Bedin, Alain Makinson, Marie-Christine Picot, Frank Mennechet, Fabrice Malergue, Amandine Pisoni, Esperance Nyiramigisha, Lise Montagnier, Karine Bollore, Ségolène Debiesse, David Morquin, Penelope Bourgoin, Nicolas Veyrenche, Constance Renault, Vincent Foulongne, Caroline Bret, Arnaud Bourdin, Vincent Le Moing, Philippe Van de Perre, Edouard Tuaillon

## Abstract

We assessed the expression of the cell adhesion molecule Sialoadhesin (CD169), a type I interferon-inducible receptor, on monocytes (mCD169) in 53 adult patients admitted to the hospital during the COVID-19 outbreak for a suspicion of SARS-CoV-2 infection. mCD169 was strongly overexpressed in 30 out of 32 (93.7%) confirmed COVID-19 cases, compared to three out of 21 (14.3%) patients for whom the diagnosis of COVID-19 was finally ruled out. mCD169 was associated with the plasma interferon alpha level and thrombocytopenia. mCD169 testing may be helpful for the rapid triage of suspected COVID-19 patients during an outbreak.

---

Like the rest of Europe, France has been severely affected by the COVID-19 epidemic. Early identification and prompt diagnosis of SARS-CoV-2 infection are of prime importance for the outcome of patients. It is also mandatory to avoid unnecessary and time-consuming interventions for patients who do not have COVID-19 but who may have another serious illness requiring appropriate care.

Type I interferons (IFN) are important factors for homeostasis of the immune response and play a key role in antiviral immunity^12^. A robust type-I IFN response is critical in the early phase of SARS-CoV-2 infection to limit SARS-CoV-2 replication and avoid a severe clinical outcome^3^. CD169, also known as sialoadhesin or Siglec-1, is constitutively expressed at low levels on monocytes, but its expression rises dramatically when monocytes become stimulated by IFNα and all other type I IFNs^1-4^. Studies have reported that monocyte CD169 (mCD169) overexpression is associated with acute viral infections^5-7^. Furthermore, a subset of CD169 lung-resident macrophages that have immunoregulatory functions and proliferate after influenza infection has recently been described^8^.

In March 2020, Montpellier University Hospital reorganized its service delivery in response to the COVID-19 outbreak. A specific care system strengthened the triage, diagnosis and hospitalization of patients with suspected COVID-19. In this prospective observational study conducted during the COVID-19 outbreak, we evaluated mCD169 expression for the identification of SARS-CoV-2 infection in patients at hospital admission.

Among 162 patients admitted in the respiratory emergency unit from 15 March to 05 April 2020, 53 (32.7%) were randomly selected and tested for mCD169 expression (Supplemental Fig.1). Thirty-two of these patients tested positive for SARS-CoV-2 RNA (31 on the first nasopharyngeal sample, one on the third sample), and 21 tested negative. Thirty patients with a confirmed diagnosis of COVID-19 had a mCD169 expression level above the positivity threshold (93.7%) (Fig.1.A). In contrast, only three of the 21 patients uninfected by SARS-CoV-2 had a mCD169 overexpression (14.3%). Neutrophil CD64 (nCD64) was assessed as a marker for systemic bacterial infection^9^. High nCD64 expression was observed in seven out of 21 (33.3%) COVID-19 negative patients (Fig.1.B). Bacterial infection was microbiologically confirmed for all but one of these patients (6/7). Four out of 32 (12.5%) confirmed COVID-19 cases also had nCD64 expression over the threshold; two of them had confirmation of a bacterial infection. A higher median expression of CD38^bright^ on CD8+ T-cells also was observed in the COVID-19 confirmed group compared to the COVID-19 uninfected group. CD38 on CD8+ T-cells is a robust marker of inflammation, overexpressed in viral infections^10^. However, this marker has a limited capacity to identified COVID-19 cases since CD38^bright^ on CD8+ T-cells was at a normal level in 11 COVID-19 confirmed cases (34.3%) (Fig.1.C). The level of mCD169 was inversely correlated with the CT (Cycle time detection) value of the SARS-CoV-2 PCR, suggesting that the immunological marker was associated with the level of virus replication (R^2^= 0.24) (Fig.1.E). At hospital admission, mCD169 was not associated with the amount of time following the onset of symptoms in COVID-19 confirmed patients (Fig.1.D). We did not observe a difference in mCD169 expression between patients who required a stay in intensive care units, patients with mild forms of COVID-19, or patients who died. The mCD169 level was correlated with the concentration of IFNα in plasma and thrombocytopenia, but not with the CRP concentration, lymphopenia or neutrophil count (Supplemental Fig. 2).

**Figure 1.**
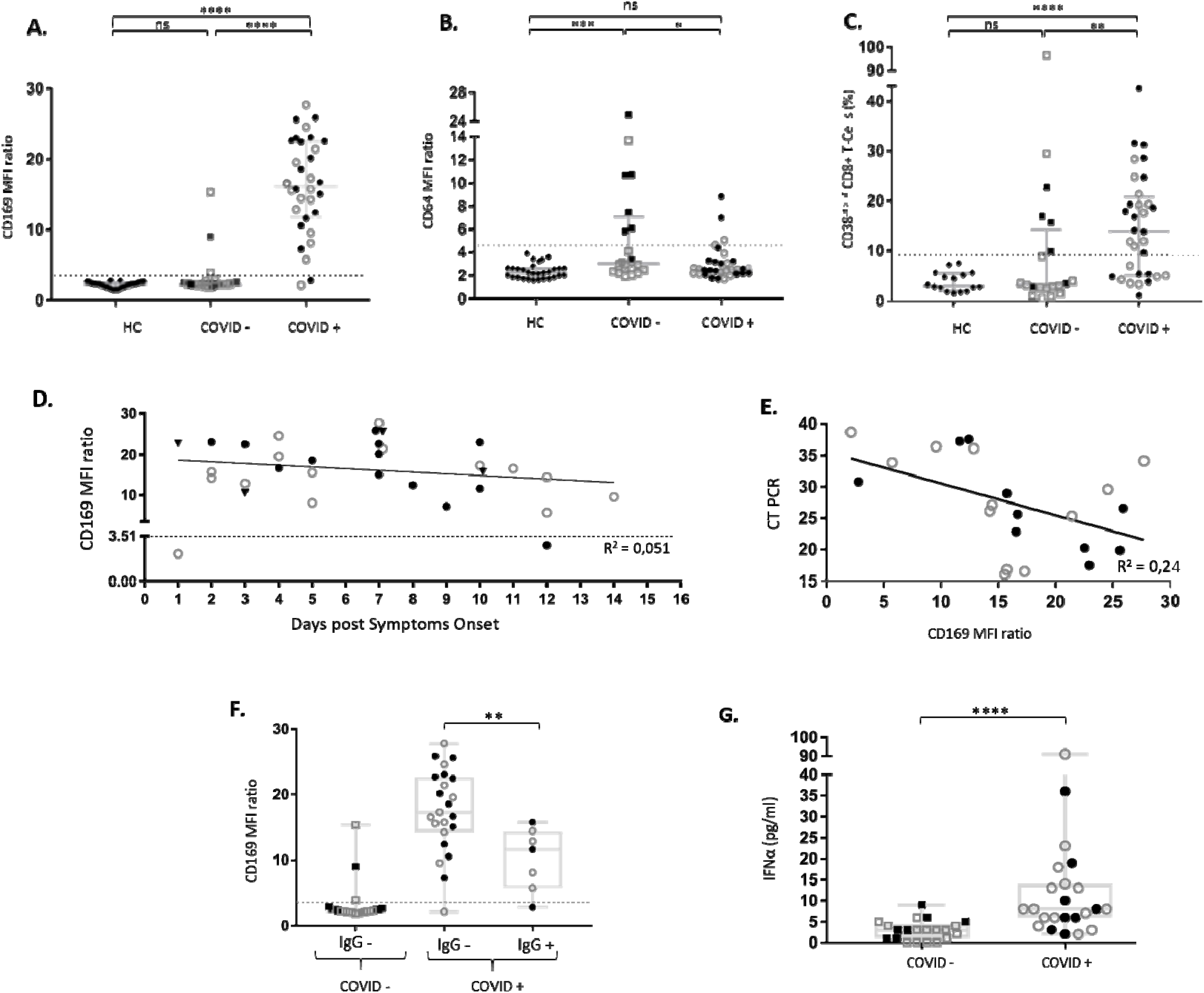
CD169 expression on monocytes in patients admitted in COVID-19 hospital units with a diagnosis of SARS-CoV-2 infection confirmed or ruled out. **A.** CD169 Median of fluorescence intensity (MFI) ratio (CD169 on monocyte / CD169 on lymphocytes). Healthy controls (HC) are indicated by black diamonds; patients tested negative for SARS-CoV-2 infection (COVID -) are indicated by open squares or black squares in patients with bacterial microbiologically confirmed infections; SARS-CoV-2 confirmed infection (COVID +) are open circles for mild and black circles for severe COVID-19. The threshold of CD169 MFI ratio is indicated by the dotted line: 3.51. ****p<0.0001; *p<0.05. **B.** CD64 MFI ratio (CD64 on neutrophil granulocyte / CD64 on lymphocytes), threshold: 4.59. **C.** Percentage of CD38^bright^ CD8+ T cells; threshold: 9.06%. **D.** CD169 MFI ratio at hospital admission according to time elapsed from the first symptoms. **E.** Correlation between CD169 MFI ratio and CT value of the SARS-CoV-2 reverse-transcriptase-real-time polymerase chain reaction (CT-PCR) in COVID-19 confirmed patients. **F.** Expression of CD169 on monocyte according to serological status for anti-SARS-CoV-2 IgG at hospital admission. **G.** Interferon alpha plasma concentration at hospital admission in patients with a diagnosis of SARS-CoV-2 infection confirmed or ruled out.

**Figure 2.**
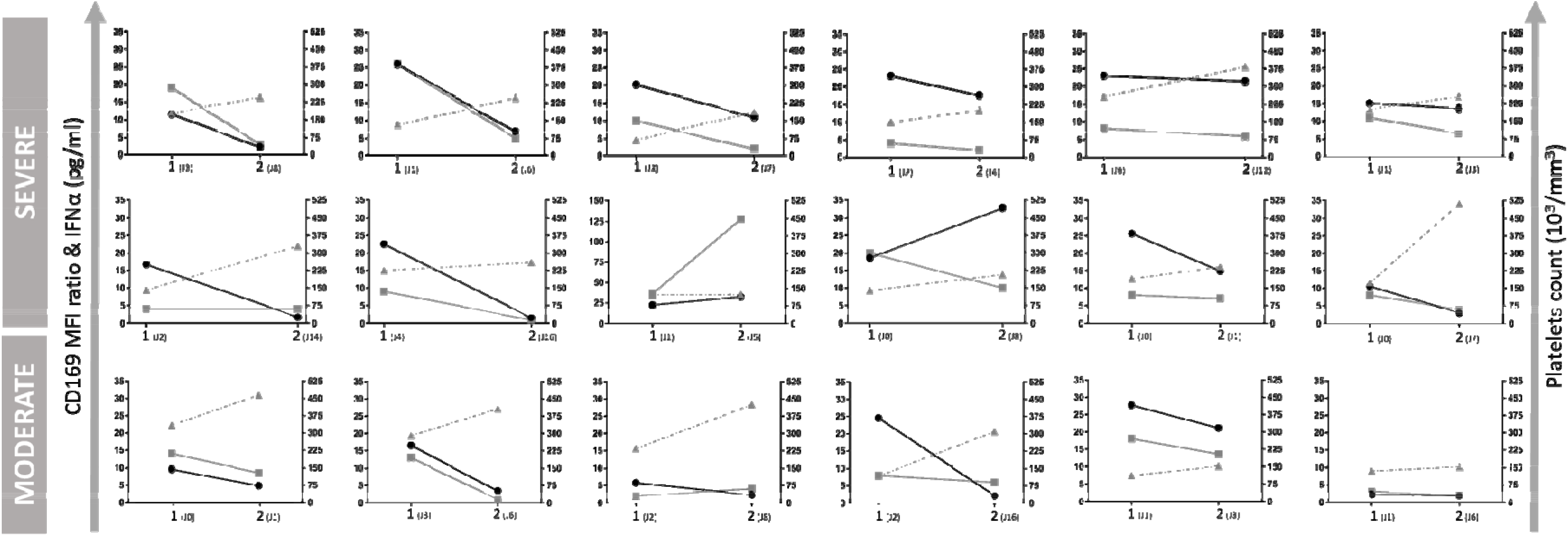
Evolution of CD169 MFI ratio, IFNα plasma concentration and platelet count on repeated testing in 18 COVID-19 confirmed patients. CD169 MFI ratio is indicated by black circles, IFNα plasma concentration by grey squares and platelets count by triangles. Days from onset of symptoms are indicated for the first and second time points.

The test accuracy of mCD169 to predict COVID-19 infection was studied by means of a receiver operating characteristic curve (ROC curve) and compared to the CRP level (Supplemental data 3). The sensitivity and specificity at the optimal operating point were 97% and 80%, respectively, with an Area under the ROC Curve (AUC) of 0.95. CRP had a sensitivity and specificity of 94% and 33%, respectively, with an AUC of 0.58. To investigate whether mCD169 could complement serological testing, we retrospectively assessed anti-SARS-CoV-2 nucleocapsid IgG. Among the confirmed COVID-19 cases, seven patients also tested positive for SARS-CoV-2 IgG at the time of hospital admission. These patients had a lower mCD169 level than patients who tested negative for anti-SARS-CoV-2 IgG (p=0.0084), suggesting that high mCD169 is associated with active SARS-CoV-2 infection, before seroconversion (Fig.1.F).

A second blood sample was taken from 18 confirmed COVID-19 patients before they were discharged from the hospital to assess changes in mCD169 levels. A concomitant decay of mCD169 and IFNα was observed in 15 out of the 18 cases (83%), with five of them achieving a normalization of mCD169 expression (Fig. 2). Two patients exhibited a rise and one a stable mCD169 expression. Of note, the platelet count increased between the first and second time points in all of the confirmed COVID-19 patients.

The rapid identification of COVID-19 is a major challenge for emergency units, especially when a hospital has to cope with 200 to 300 suspected cases but a low number of real infections. Montpellier University Hospital has been strongly impacted by the COVID-19 crisis although the region around Montpellier reported a relatively low proportion of COVID-19 cases during the peak of the epidemic, which occurred in France between March and April. This was due to the dynamics of the spread of the disease in France. The first cases of SARS-CoV-2 were reported in the country in mid-February 2020^11^. The spread of the virus accelerated thereafter, in particular from a large, four-day long religious meeting that began on February 17 in a city located 650 km from our hospital. Since France went into lockdown on 17 March 2020, the COVID-19 burden in the Montpellier region has been very different compared to the harder hit northeastern part of the country. The seroprevalence (95% CI) at this end of the wave^12^ was estimated at 1.9% (1.2-3.3) in the Montpellier region versus 9.1% (6.0-14.6) in the northeast. Consequently, even at the peak incidence of SARS-CoV-2 (when the study was conducted), only some of the patients suspected of COVID-19 and admitted in dedicated care units had a confirmed diagnosis of SARS-CoV-2 infection. For the remainder, the diagnosis of COVID-19 needed to be rapidly ruled out following hospital admission. To our knowledge, the present study is the first to evaluate the value of mCD169 measurements in the rapid identification of COVID-19. We found that mCD169 could predict SARS-CoV-2 infection at hospital admission during the outbreak. This biomarker could be relevant for triage and rapid therapeutic decision in patients suspected of acute viral infections since mCD169 can be overexpressed in other infections responsible of outbreaks such as influenza or dengue fever.

Our study has some limitations. First, we did not included subjects with negative PCR results but later COVID-19 confirmation by radiological or serological findings. Second, the small number of subjects did not allow us to explore the relationship between the dynamic changes of mCD169 and patients’ outcomes. Finally, we could not confirm our findings through a large-scale survey due to the rapid decrease in admissions of suspected COVID-19 cases in our hospital after April.

In conclusion, mCD169 may be a sensitive marker for the rapid triage of patients suspected of having COVID-19. This type I IFN-inducible receptor is strongly overexpressed on monocytes during the early phase of the SARS-CoV-2 infection, and remains elevated in patients admitted to the hospital during the second week following the onset of symptoms. The assessment of mCD169 may complement viral and serological methods to improve the diagnosis of COVID-19. Alongside RT-PCR and serological testing, mCD169 may contribute to preserving the medical capacities of emergency departments by favoring the rapid orientation of patients with possible COVID-19. The value of leukocytes activation markers, including mCD169 and nCD64, in the diagnosis of acute infection needs to be evaluated during viral outbreaks in clinical studies. The development of fully automated tests for these markers may be crucial to prepare for future potential epidemics.

## Material and methods

This study was conducted by the University Hospital of Montpellier, the 7^th^ largest hospital in France, during the peak of the COVID-19 epidemic between 15 March and 5 April 2020. The population of this prospective, observational study consisted of consecutive adult patients suspected of SARS-CoV-2 infection and hospitalized in the medical units dedicated to the diagnosis of COVID-19. There was a limit of 15 patients enrolled per day due to the limited capacity of the laboratory to perform cytometer analysis during the outbreak. The patients satisfied the criteria of the national guidelines for RT-PCR testing, which at the time of the study consisted of the presence of symptoms of pneumonia or having risk factors associated with severe COVID-19. Cases for whom the diagnosis of COVID-19 could not be confirmed or dismissed based on RT-PCR results were excluded from the study. Of the 52 patients recruited, 32 patients were confirmed to be SARS-CoV-2 positive using a deep throat-swab, while 21 tested negative. Among the confirmed cases, 18 patients had a second cytometry analysis during their hospitalization. The median age of the patients was 64 years (IQR, 51–78.5 years) and 54.7% were male (Supplementary Table 1).

### Positive controls and thresholds

We assessed the expression of mCD169, nCD64 and CD38^bright^ on CD8 T-cells in 30 healthy controls to establish a threshold based on the median + 3SD. The thresholds were: 3.51 for the mCD169 MFI ratio, 4.59 for the nCD64 MFI, and 9.06% for CD38^bright^. The nCD64 threshold was controlled on 10 clinical samples collected from patients with microbiologically confirmed bacterial infections (median (IQR) = 8.44; (5.77-11.37)).

### SARS-CoV-2 RNA testing

SARS-CoV-2 RNA extraction from nasopharyngeal swabs (Sigma Virocult, Medical Wire Instrument, Corsham, UK) was done using the QIAamp Viral RNA Mini Kit on the QIAsymphony platform, following the manufacturer’s instructions. SARS-CoV-2 RNA was assessed using a RT-PCR targeting RNA-dependent RNA polymerase (RdRp) as previously described^13^.

### Flow cytometry

10 µl of EDTA sample was simultaneously lysed with 500 μL of Versalyse lysing solution and stained with CD64-CD169/infections dried custom mixture, composed of anti-CD169-phycoerythrin (PE) (clone 7-239) and anti-CD64-Pacific Blue (PB) (clone 22). A second panel was used containing CD45-FITC, CD8-ECD, CD3 PC5, CD4-RD1 (CYTO-STAT tetraCHROME) and CD38-PB. Samples were incubated at room temperature for 30 minutes, in the dark. All products or custom products came from Beckman Coulter Inc, (Brea, CA). We acquired on a 3-laser, 10-color Navios flow cytometer and analyzed using Kaluza Software version 2.1 (both from Beckman Coulter Brea, CA Inc). The stability of the instrument was monitored and validated daily with Flow-Check Fluorospheres used to verify instrument optical alignment and fluidics, and Flow-Set Fluorospheres for the quantitative analysis of human leukocytes.

### Interferon alpha plasma concentration

Plasma samples were stored at − 80 °C until processing. Interferon alpha (IFN-α) was quantified using a multiplex microsphere assay (Invitrogen Human Inflammation 20-plex ProcartaPlex Panel, Marne-La-Vallée, France) on a Luminex apparatus (MAGPIX, Thermo Fisher Scientific, Massachusetts, USA) following manufacturer’s instructions.

### Serology

Plasma samples were tested for IgG antibodies directed against SARS-CoV-2 nucleocapsid using a CE-marked ELISA (ID.Vet, ID screen® SARS-CoV-2-N, Montpellier, France) as previously described^11^. Each sample is treated as a ratio: S/P (Sample/positive control) expressed in percentile (%): S/P % = (Optic density (OD) of sample – OD negative control) / OD positive control – OD negative control) x 100. S/P > 60% is positive and < 60% is negative.

## Statistical analysis

Data were analysed and illustrated using Excel 2016 (Microsoft Corp, Redmond, Washington) and Prism 7 (GraphPad Software Inc, La Jolla, California) software. To determine statistical significance between the two groups (ie. CD169 COVID+ vs COVID -), unpaired Student’s two-tailed t-test or non-parametric Mann-Whitney test was applied, according to the distribution. Correlations were analysed using Pearson or Spearman’s rank test according of the normality of the data set. To determine statistical significance between two groups (i.e., CD169 COVID + versus COVID -), an unpaired Student’s two-tailed t-test was applied.

A p value of <0.05 was considered statistically significant.

## Data Availability

Data will be obtainable after acceptance of the manuscript for publication through a simple request to the corresponding author.

## Acknowledgements

This work was supported by the Montpellier University Hospital, Muse I-SITE Program grant, University of Montpellier. The funders had no role in the design of the study, the collection or analysis of data or the decision to publish. We thank S. Groc, S. Marie, and C. Niel for technical assistance.

## Contributions

A.S.B. planned and supervised laboratory testing, analyzed the data and wrote the manuscript. A.M, D.M, A.B, V.L.M managed patients and evaluated clinical data. A.P, L.M., E.M, K.B, S.D., V.F, and C.B performed laboratory testing. F.M wrote the manuscript. M.C.P. evaluated clinical data and supervised statistical analysis. P.Vd.P. designed the study and wrote the manuscript. E.T. designed and conceptualized the study, analyzed the data and wrote the manuscript.

## Ethical approval statement

All patients were included in the COVIDOtheque cohort (ClinicalTrials.gov Identifier: NCT04347850) and provided informed consent for the use of their data and clinical samples for the study. Institutional review board clearance for the scientific use of patient data has been granted by the Institutional of Montpellier University Hospital and Ile de France III ethical committee (n°2020-A00935-34).

